# A Novel Heuristic Global Algorithm to Predict the COVID-19 Pandemic Trend

**DOI:** 10.1101/2020.04.16.20068445

**Authors:** Panagiotis G. Asteris, Maria Douvika, Christina Karamani, Athanasia Skentou, Tryfon Daras, Liborio Cavaleri, Danial Jahed Armaghani, Katerina Chlichlia, Theoklis E. Zaoutis

## Abstract

Mathematical models are useful tools to predict the course of an epidemic. A heuristic global Gaussian-function-based algorithm for predicting the COVID-19 pandemic trend is proposed for estimating how the temporal evolution of the pandemic develops by predicting daily COVID-19 deaths, for up to 10 days, starting with the day the prediction is made. The validity of the proposed heuristic global algorithm was tested in the case of China (at different temporal stages of the pandemic). The algorithm was used to obtain predictions in six different locations: California, New York, Iran, Sweden, the United Kingdom, and the entire United States, and in all cases the prediction was confirmed. Our findings show that this algorithm offers a robust and reliable method for revealing the temporal dynamics and disease trends of SARS-CoV-2, and could be a useful tool for the relevant authorities in settings worldwide.

## Introduction

In January 2020, the novel coronavirus SARS-CoV-2 was identified as the causative agent of an outbreak of viral pneumonia known as coronavirus disease 2019 (COVID-19) in Wuhan, China. As of 1 May 2020, this outbreak of COVID-19 has spread to more than 200 countries and has been officially declared a global pandemic [1]. The number of confirmed cases and deaths continues to increase every day. As of May 4, there have been 3,47 million cases and 246, 979 deaths confirmed worldwide.

In response to COVID-19, governments have implemented mitigation measures to reduce density, measures which have also affected their economies; approximately 3 billion people are under lockdown worldwide as of early May 2020. Mathematical models are used to forecast the course of the epidemic and guide governments and national authorities in making key decisions for rapid strengthening of outbreak surveillance and management. However, there is an urgent need for tools capable of early prediction, in order to further bolster the accuracy and efficiency of government measures [2]. It has been shown that short-term forecasts can guide the intensity and type of interventions needed to mitigate an epidemic [3-5]. In the light of the above, the aim of the present study was to apply a novel robust and reliable global algorithm for estimating and predicting the COVID-19 pandemic trend for up to 10 days starting with the prediction date in two U.S. states (California and New York) and four countries (Iran, Sweden, UK, USA). In the case of USA, the algorithm was applied both to these two states as well as to the country as a whole, in order to investigate how the pandemic is developing in different parts of the same country.

## Methods

### Assumptions and Data sources

During the study of the development of the COVID-19 pandemic, the daily number of confirmed deaths due to COVID-19 for each location have been recorded and analyzed further. The selection of daily deaths was based on the authors’ assumption that mortality rates provide more accurate and reliable data compared to the recordings of the number of daily infected individuals. The daily mortality rate is suggestive of additional information about the unique characteristics of each setting, which influence the pandemic transmission trend in each place. Such characteristics include:

- The climate and environmental conditions in each location
- The quality of the healthcare systems in each location
- The experience and expertise of the medical staff and healthcare workers
- The age distribution of the population
- The pandemic mitigation measures applied in each location

The main assumption during the design of this algorithm was the observation that the mortality rate, in particular the death numbers in the respective populations, follow a normal distribution. Although daily recording might not be the case for optimal normal distribution, it is important to note that the selection of death recordings every two or three days almost always leads to an optimal normal distribution. Following this assumption, the simulation of the pandemic spread was investigated for a variety of mortality rates in different settings, and the setting giving the most accurate results and predictions was selected in developing the model.

In the process of developing a new prediction model, it is common that scientists pay attention to the computational model; yet a reliable database is of high importance, and in order to achieve a reliable forecast, researchers should give the appropriate attention to the database used for the development, training, and validation of the model. In the light of the above, the final overall database was based on two individual databases. Data for the countries were obtained from the database Worldometer [6] and, for the United States, from the COVID Tracking project [7]. In our prediction we did not accommodate underreporting of cases or deaths, which is common in many parts of the world with considerable influence on the prediction results. Recent analysis shows that the official global COVID-19 death toll is much higher (60%) than officially reported [8].

### Proposed Heuristic Algorithm

By analyzing the official data from China, including daily COVID-19 infections and deaths, it is clear that they can be expressed with meaningful accuracy using a suitable Gaussian curve (or, equivalently, a proper normal distribution density function). In addition, by studying the evolution of the pandemic and the course of the restrictions in this country, and taking into account that many European and other world countries have taken similarly strict restrictive mitigation measures, we assumed that the development of COVID-19 pandemic would have similarities to its development in China. In other words, we propose that the number of new incidents or deaths will be expressed using a proper normal distribution.

A Gaussian function is a function of the form: 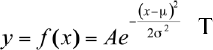, whose graph is a symmetrical bell-shaped curve centered at the position *x* = µ · *A* is the height of the peak and the variance σ^2^ controls its width. On both sides of the peak, the tails of the curve quickly fall of and approach the x-axis (asymptote). Our algorithm aims to determine in each state or setting the optimal normal curve for daily deaths by calculating the parameters *A* µ σ^2^; i.e. by fitting the “best” possible normal curve. The optimality of the normal curve is given with reference to well-known statistical indices.

More precisely, the main steps of the algorithm are (through a triple loop):

1. (for A / first inner loop) We start from a given value of A, and with step 1, we continue up to a certain value (desired accuracy) depending on the maximum value of our available data (deaths)
2. (for σ^2^ / second inner loop) 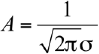 so peaking A, σ^2^ can be calculated. The algorithm then uses a probability value p, starting from p=0.85, and with step 0.01, continues up to 0.99999 (it is known that *P* = *px*100% of the data under a normal distribution curve lie inside the interval 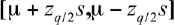, where *q* = 1 − *p* and 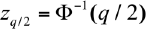, Φ the cumulative distribution function of the standard normal distribution (*N*0,1) and Φ^−1^ its inverse function. This interval is used to fit the actual data, using a proper transformation).
3. (for μ / third inner loop) We start from a value of μ =20 and we continue, with step 1 (day), up to a value of μ=60 (we observed for example that in the case of China, the phenomenon lasted for about 60 days with an average [peak day of deaths] at about the 30th day).
4. The algorithm application creates a large number of proper normal distributions by calculating in each of the three parameters (theoretical/experimental values) each time. Finally, these values are compared with the empirical values (actual numbers of deaths) and the “best” possible curve is being selected using a number of indices, i.e. the algorithm searches for the optimal curve characteristics using the available data up to the forecast day.

### Performance Assessment

The reliability and accuracy of the best fit Gaussian curves developed for each one prediction were evaluated using Pearson’s correlation coefficient R and the root mean square error (RMSE). RMSE presents information on short-term efficiency, which is a benchmark of the difference in predicated values compared to the experimental values. A lower RMSE indicates a more accurate evaluation. The Pearson’s correlation coefficient R measures the variance that is interpreted by the model, which is the reduction of variance when using the model. R values ranges from 0 to 1, with the model having the healthiest predictive ability when it is near to 1 producing little analysis when it is near to 0. These performance metrics are a good measure of the overall predictive accuracy.

### Methodology

The present section outlines the methodology used to investigate the spread of COVID-19 in a state, city, region, or country. As an example, the methodology is presented here step by step, as it was conducted and applied in the case of the investigation of the spread of the epidemic in China. Given that the outbreak in China preceded the spread of the epidemic to other countries, this allows us to apply the proposed algorithm at the beginning of the phenomenon; in the intermediate phase, which is usually characterized by a strong dynamic; and at its peak, where the phenomenon begins to fade or recede.

The main principles of the proposed methodology are as follows:

- In each step, the optimal normal distribution is calculated using the proposed algorithm and based on data available at the moment of calculation.
- The first assessment is made 14 days after the first death record. The period of two weeks is considered necessary to reliably characterize the beginning of the phenomenon At each step, following the 14-day period from the first death recorded, the optimal data simulation curve is calculated with the use of the proposed algorithm. Figure 1 shows the optimal curves for the country of China that best simulate the number of deaths for three different days (6, 12, and 18 February 2020).
- The same procedure is applied on a daily basis. Based on the values of the maximum number of deaths and on the time when this maximum is attained, the curve shown in Figure 2 is plotted. This figure illustrates how the phenomenon evolves by providing us with an estimate of when the phenomenon, including the number of deaths, is expected to peak. This information is helps authorities prepare and make informed decisions about mitigation and containment measures.
- By predicting the optimal curve of Figure 1, we are also provided with information about the dynamics of the phenomenon. Specifically, by knowing the parameters of the distribution (sigma, mi, and fitting probability), its area can be calculated, which shows the total predicted number of deaths. Based on the percentage change in the number of deaths as a function of time, the change in the dynamics of the phenomenon is defined (Figure 3). This figure strongly demonstrates that the COVID-19 phenomenon is a predominantly dynamic phenomenon with clear dynamic characteristics which oscillates strongly during its transition to the peak, and then dissipates where the balance of the pandemic phenomenon takes place. This diagram is can be used to quantify the effectiveness of the control and mitigation measures starting from the first day of application, and it provides a temporal prediction of the time period required for the dampening of the phenomenon.
- In addition to the producing the above estimates and revealing the dynamic characteristics of the phenomenon, the proposed heuristic algorithm also enables reliable prediction of the expected number of deaths for the next 10 days (Figure 4). The algorithm provides simultaneous estimates for its higher and lower limits. Based on a comprehensive study in the six aforementioned locations and the results presented below, these limits, as well as the difference between the predicted and actual deaths, were confirmed for all locations.

**Figure 1.**
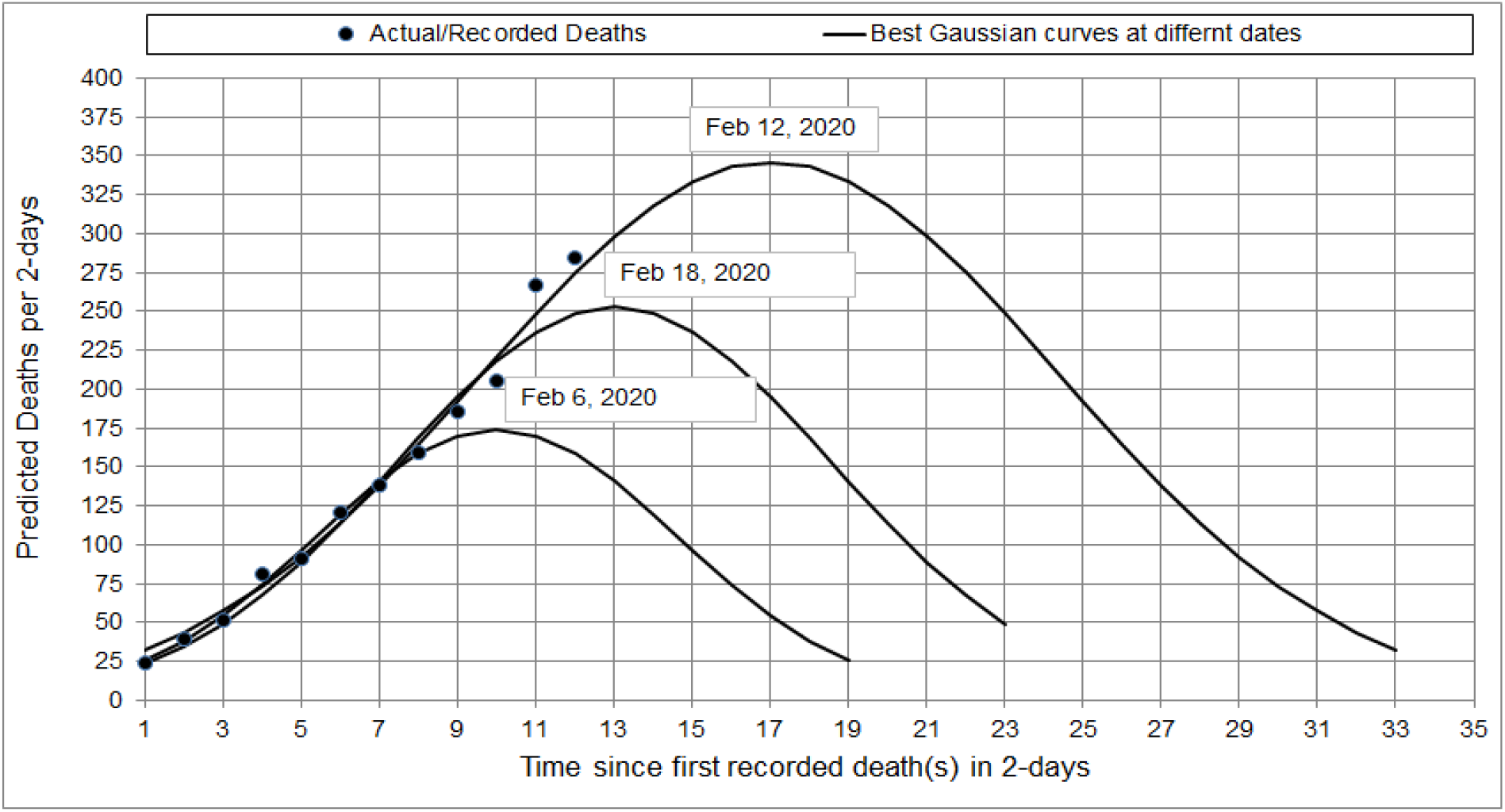
Prediction of the best Fit curves of the actual deaths at three different dates of prediction (Country of China)

**Figure 2.**
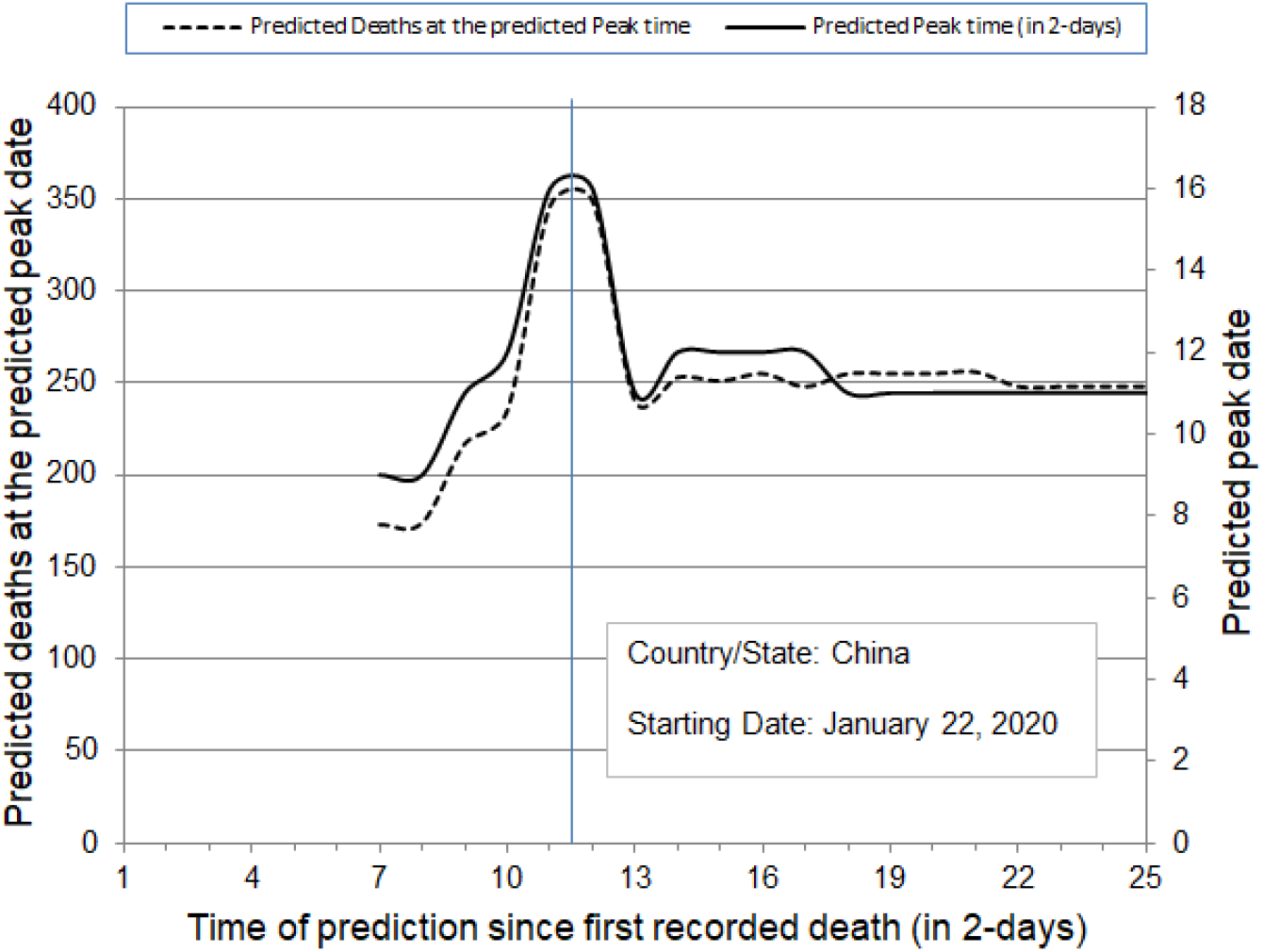
Prediction of number of deaths (dashed line), in 2-day time intervals, and peak date of deaths (solid line) in China

**Figure 3.**
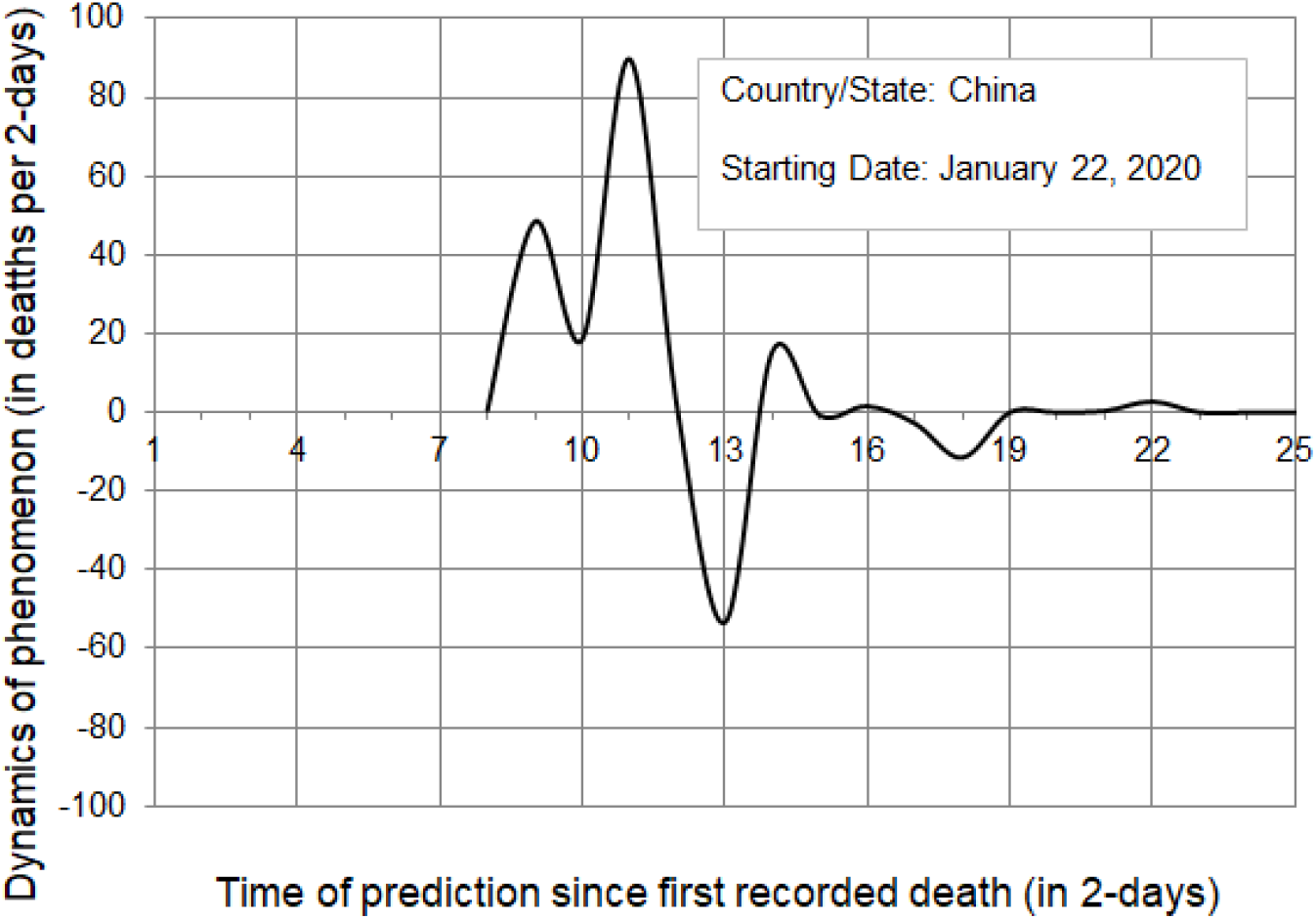
Prediction of the dynamics of COVID-19 phenomenon for the country of China

**Figure 4.**
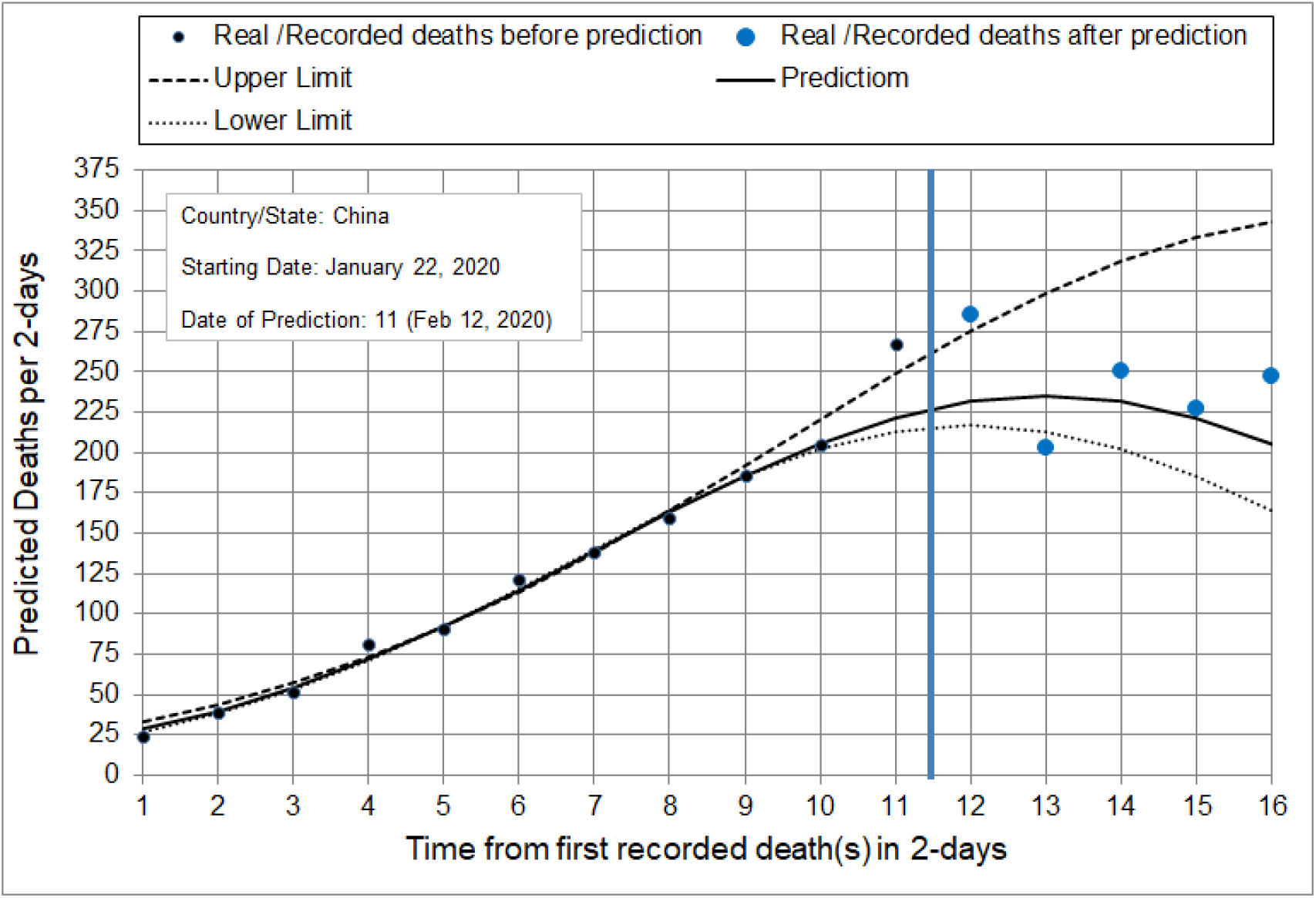
Prediction of number of deaths in China, in 2-day intervals, for the next ten days starting 12 February 2020. Black dots represent actual data until the day on which the algorithm made the prediction. Blue dots represent actual data after the day on which the algorithm made the prediction.

## Results and Discussion

In order to predict the course of the COVID-19 epidemic, a new computer software has been developed at the Computational Mechanics Laboratory, School of Pedagogical and Technological Education, Athens, Greece. Utilizing this software through implementation of the heuristic algorithm, the development of the epidemic was investigated in six different geographical locations: the states California and New York, the United States as a whole, and Iran, Sweden, and the United Kingdom. The investigation was implemented in two stages. In the first stage the data as well as the daily mortality rates and results were known. This stage served to evaluate the phenomenon, as well as to document the proposed heuristic global algorithm and its methodology. In the second stage, predictions were made for which the results are unknown. For all the above-mentioned locations the number of daily deaths was predicted for the 10 consecutive days, following 13 April 2020. The results are presented in detail for each location in the tables and figures in the supplementary materials.

The main results of our study are the following:

- In the first phase, where data and results of daily mortality rates were known, the proposed algorithm was confirmed in all the cases where it was implemented.
- The proposed methodology provides an upper and a lower estimation limit, which was confirmed for the total cases in the first stage.
- For the secondary stage, and specifically for the time period 13 - 22 April, the predictions are in Table 1. For each setting, the predictions for the next ten days as well as the predicted time and the number of deaths at the predicted time are presented in the Figures in the supplementary materials. In Figure 5, the predictions for the state of California are presented (blue dots). In this figure, the number of deaths, in 2-day intervals, for the next ten days is presented, starting 13 April 2020. Black line shows the course of the prediction. Black dots represent actual data until the day in which the algorithm made the prediction, while blue dots represent the predicted data after the day in which the algorithm made the prediction. These were performed on 12 April, the same day of the current study. Please note that the upper and lower estimation limit is in our predictions much smaller than the respective limits provided by IHME as evident in Table 1. We have already uploaded our predictions (April 16, 2020) as a preprint in MedRxiv [9] before the end of the forecasting (April 22). It is of utmost importance that all our predictions were confirmed after the end of predictions: all graphs followed the expected course (black line) as documented in each figure presenting the values for each country.
- The diagrams that present the development of the phenomenon provide additional information to support the efficacy and efficiency of the restrictive measures, including lockdown, implemented by governments to address the pandemic. A typical example is the case of China, where the impact of the dynamic diminishes 23 days after the implementation of restrictive measures. This is in accordance with other publications, where it has been suggested that there is a two- to three-week lag between the introduction of interventions and the manifestation of their impact on in hospitalized case numbers [10].

**Figure 5.**
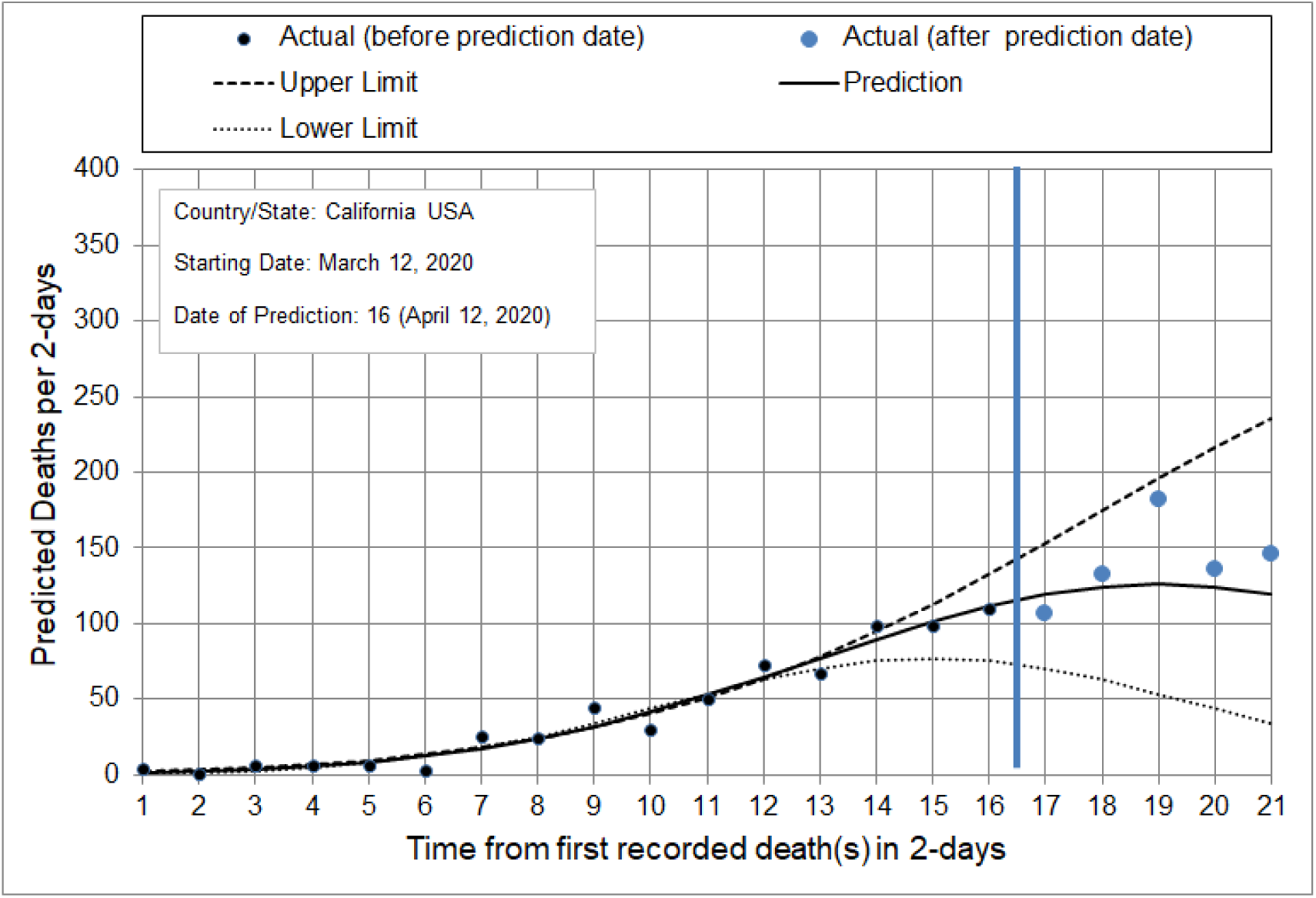
Prediction of number of deaths, in 2-day intervals, for the next ten days starting 13 April 2020, for the state of California. Black dots represent actual data until the day on which the algorithm made the prediction. Blue dots represent actual data after the day on which the algorithm made the prediction. This figure (as well as all the similar Figures presented in supplementary materials) and the predictions have been uploaded (April 16, 2020) by the authors as a preprint in MedRxiv [**9]**

**Table 1.**
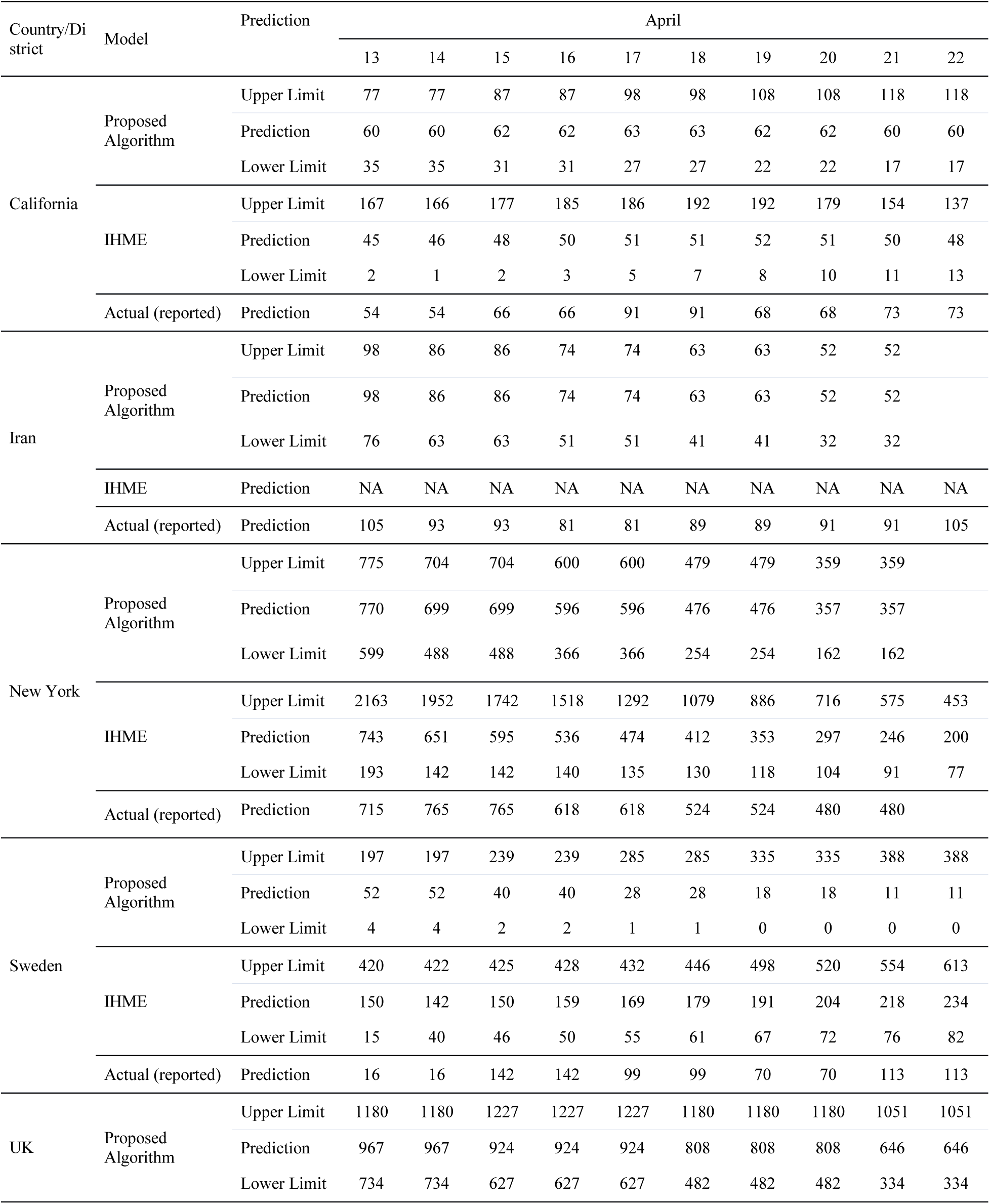
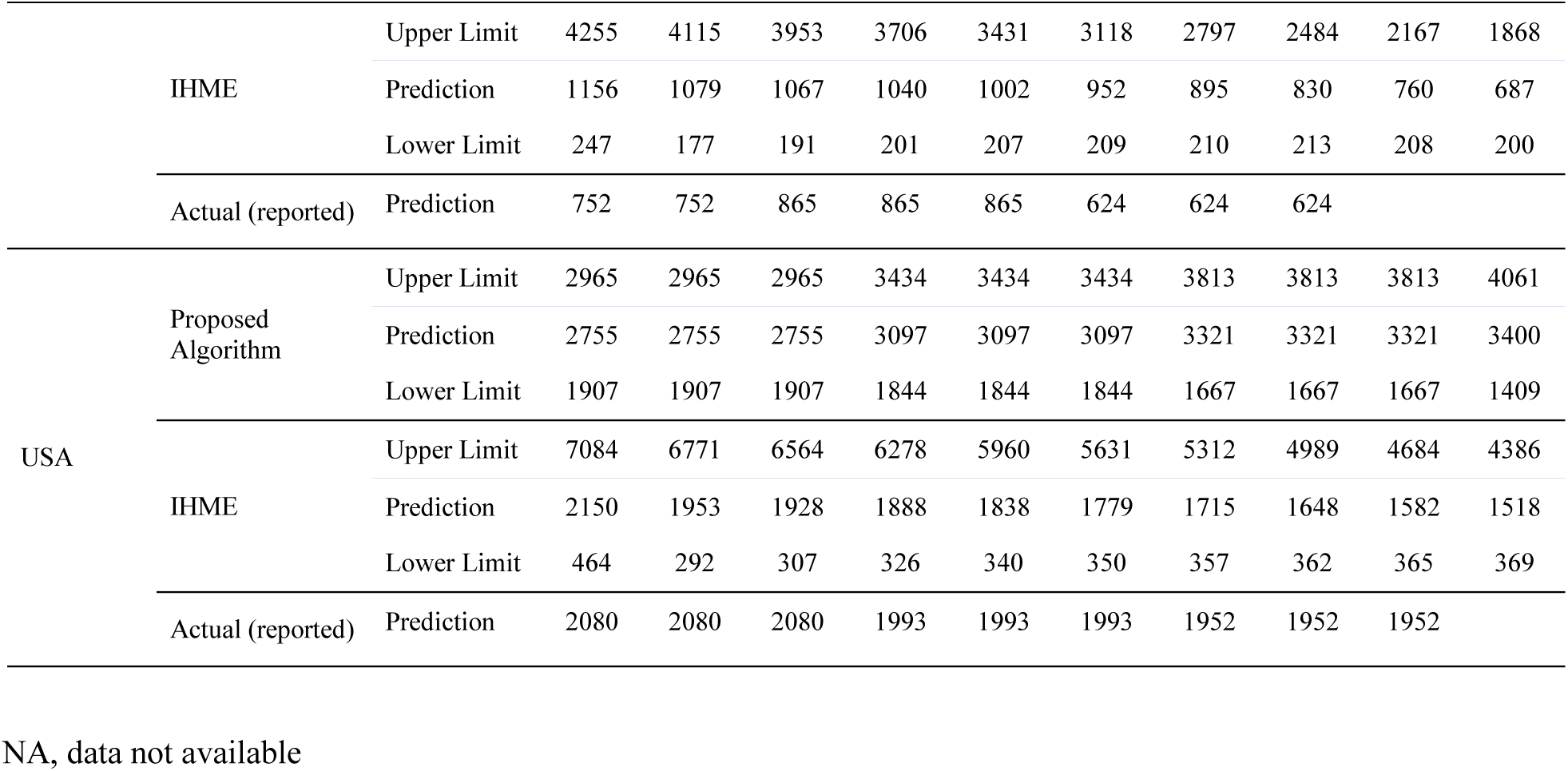
Predicted daily deaths, in the time interval 13-22 April 2020, obtained using our model compared, with corresponding predictions by the Institute for Health Metrics and Evaluation (IHME). Predicted number of deaths (estimated) refers to the number reported before 13 April 13. Actual (reported) refers to number of deaths reported after the time interval, after 22 April.

Although so far there are reports available on country-specific models [4, 11] that take into consideration the specific features of each country or region, this novel global algorithm is of particular importance as it applies to six different locations with singular and distinctive characteristics. Iran is one of the most populous countries in Eurasia, and has experienced significant infections and death from COVID-19 compared to its nearby neighbors, such as Iraq. Sweden is a Nordic country with the world’s eleventh-highest per capita income and a high ranking among countries for quality of life; it is also one of the few countries worldwide that has not imposed a lockdown or even school closures. The United Kingdom is a sovereign country northwest of Europe with a high-income GDP and nationalized healthcare, while the USA, located in North America, is the third most populous country in the world and has an unusual public-private healthcare system.

Although both New York and California are densely populated, COVID-related outcomes have been enormously different, with California experiencing a milder outbreak and New York experiencing a severe outbreak: as of 13 April, California had more than 23,000 confirmed cases and about 680 confirmed deaths, while New York State has more than 190,000 confirmed cases and about 9,400 confirmed deaths. This may be attributed to a variety of factors, from socioeconomic factors to divergences in the states’ respective management of the pandemic. Perhaps most significant is that California imposed restrictive measures earlier than New York: California reported its first death on 4 March and imposed lockdown measures starting on 19 March, while New York reported its first death on 14 March and imposed lockdown measures starting on 23 March. This shows the importance of proactive measures and the significance of available accurate predictive models.

While the states across the USA imposed localized lockdowns, the UK went into full lockdown on 23 March and Iran on 19 March. Sweden is one of the few countries that used a different strategy and did not mandate strict social distancing measures. Notably, in Figure 3, the effect of the applied measures is shown and presents the dynamic nature. Mitigation measures start to show effects three weeks after the start of the quarantine measures, and the effect is obvious in the relevant graph (Figure 3). The number of deaths slowed down 23 days after the start of mitigation interventions.

Although there are significant differences and unique local transmission dynamics among different settings, we managed to develop and apply an algorithm that is able to predict with high confidence the outcome of the outbreak in all examined states for the consecutive 10 days. Interestingly, previous reports show that good probabilistic calibrations of their forecast models was achievable also at short time horizons of one or two weeks [3-5]. Thus, despite the fact that every setting has unique internal socioeconomic and political characteristics, as well as differing policy responses to the pandemic, as a result of which the impact of the epidemic may not be evenly distributed, the global prediction tool that we have developed can be applied to all the settings tested.

## Conclusions

We applied a novel algorithm for comparatively estimating and predicting the COVID-19 pandemic trend in 6 locations for up to 10 days, starting at the prediction date. This novel global algorithm accurately predicted the outcome of the outbreak in all geographic locations in which it was applied, despite differences among the six settings and in their responses to COVID-19. The proposed algorithm may provide a useful tool for policymakers in addressing COVID-19-related preparedness and planning.

## Data Availability

Data referred to the manuscript are available

## Supplementary Materials

**Figure 1.**
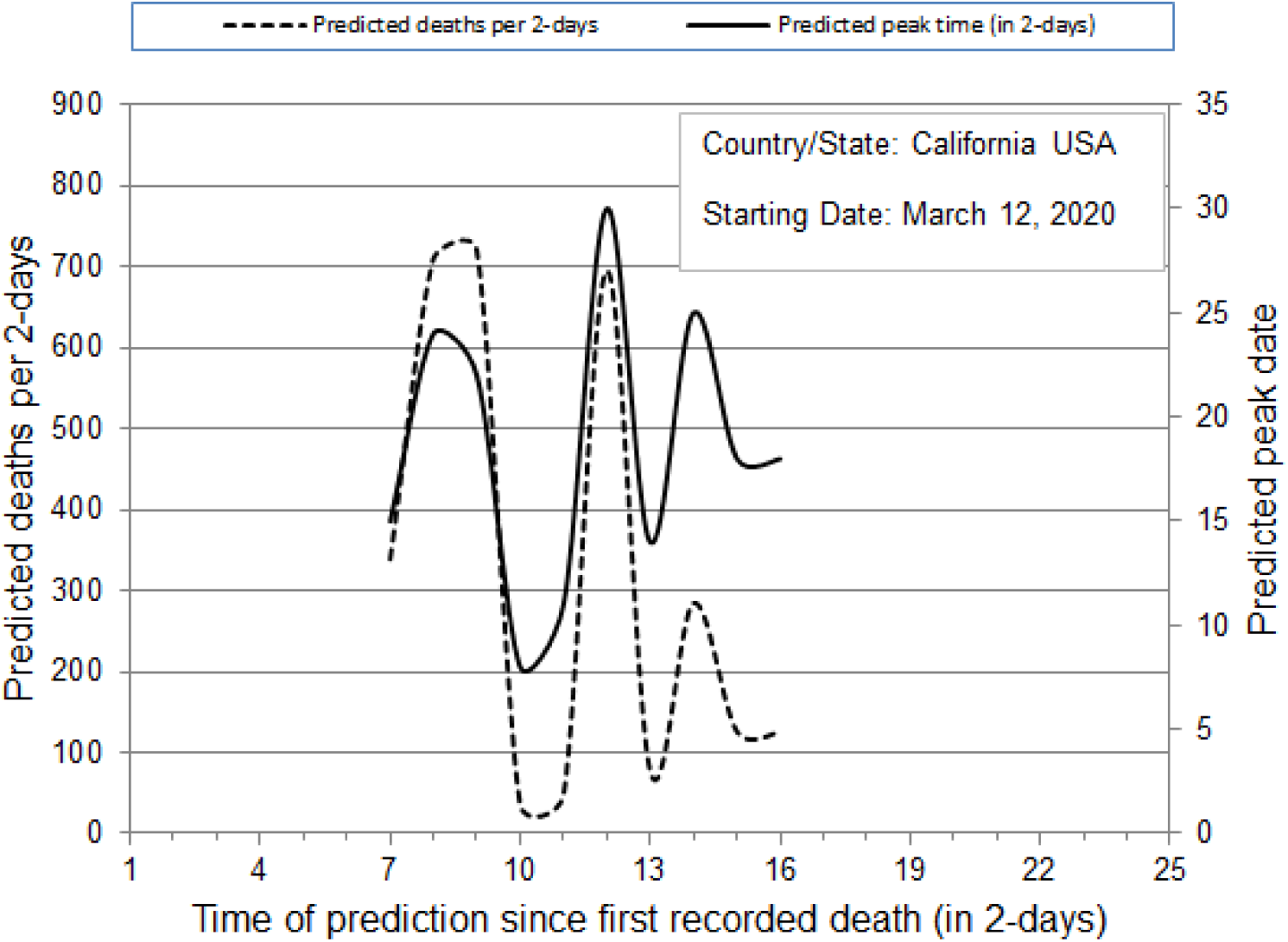
Prediction of number of deaths (dashed line), in 2-day time intervals, and peak-date of deaths (solid line) for the state of California, USA

**Figure 2.**
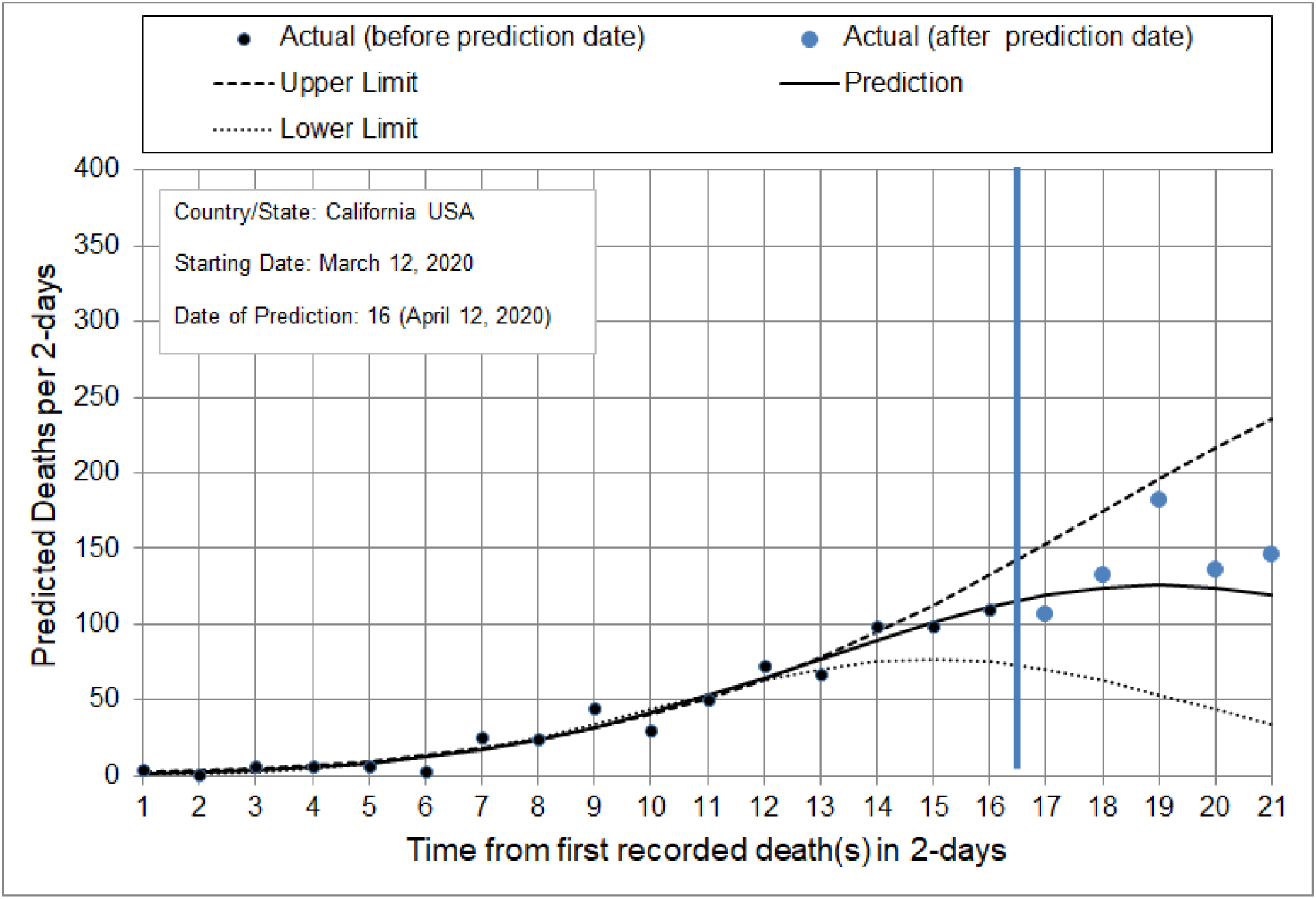
Prediction of number of deaths, in 2-day intervals, for the next ten days starting April 13, 2020, for the state of California, USA. Black dots represent actual data until the day in which the algorithm made the prediction. Blue dots represent actual data after the day in which the algorithm made the prediction.

**Figure 3.**
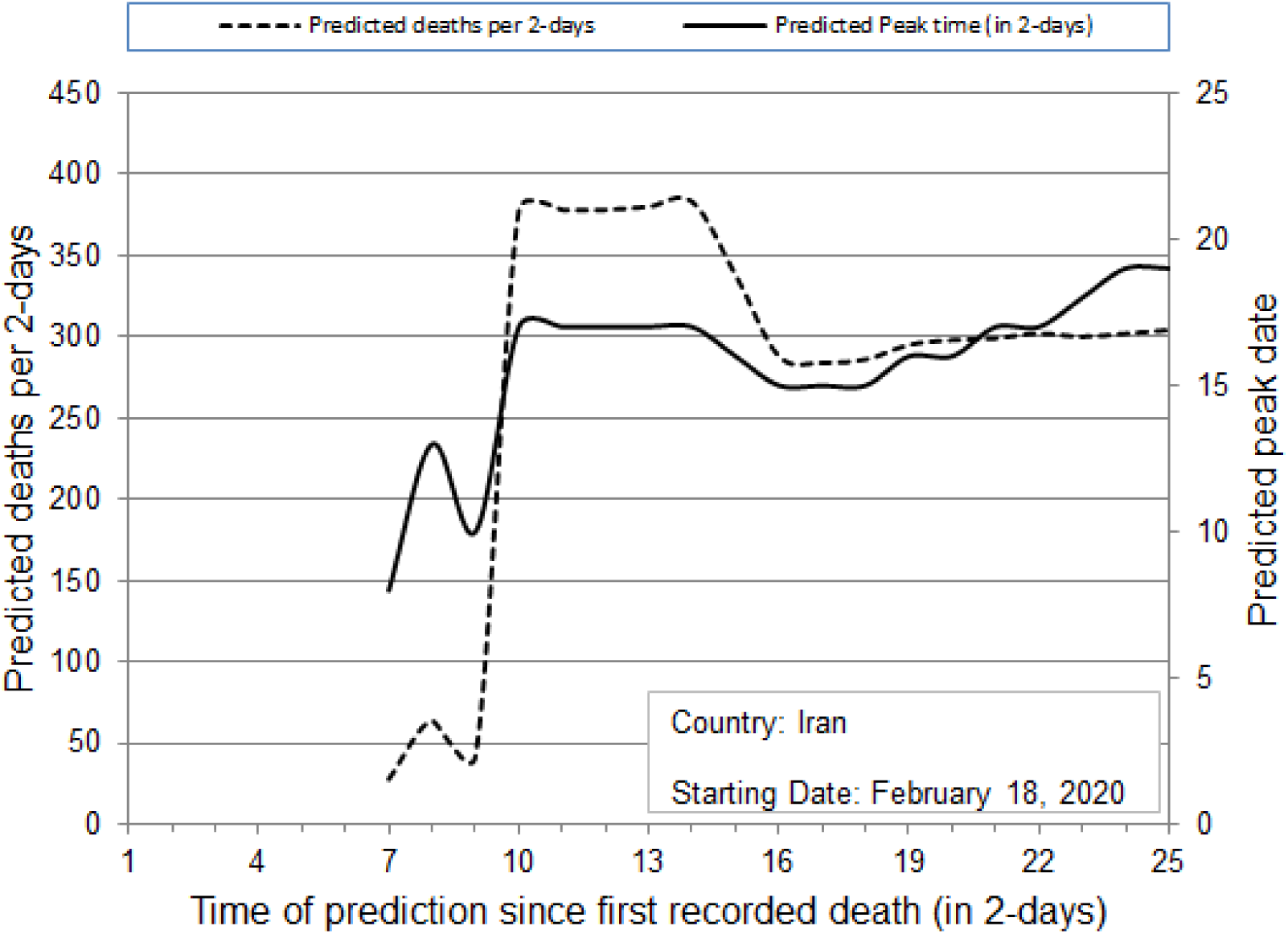
Prediction of number of deaths (dashed line), in 2-day time intervals, and peak-date of deaths (solid line) for the country of Iran

**Figure 4.**
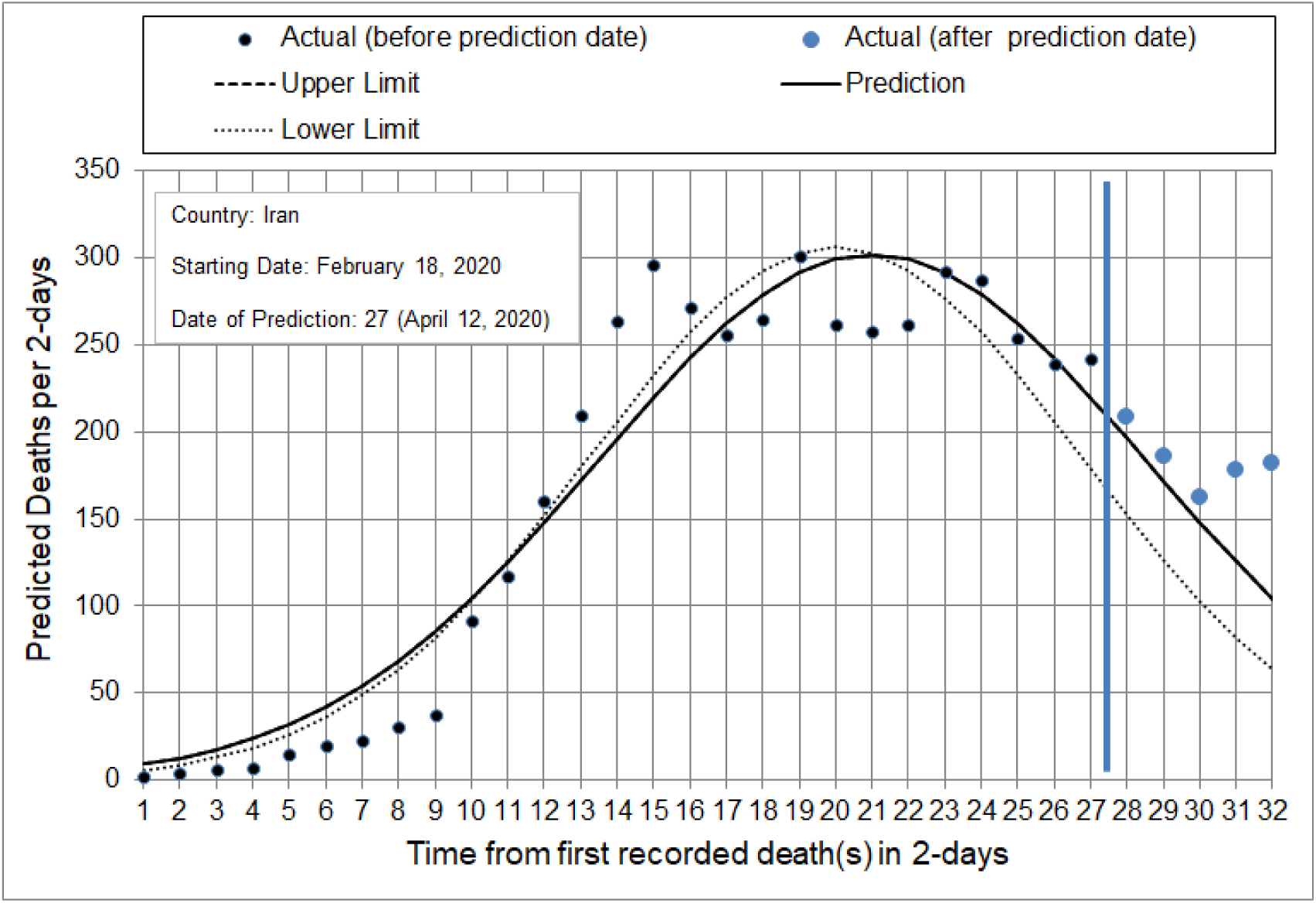
Prediction of number of deaths, in 2-day intervals, for the next ten days starting April 13, 2020, for the country of Iran. Black dots represent actual data until the day in which the algorithm made the prediction. Blue dots represent actual data after the day in which the algorithm made the prediction.

**Figure 5.**
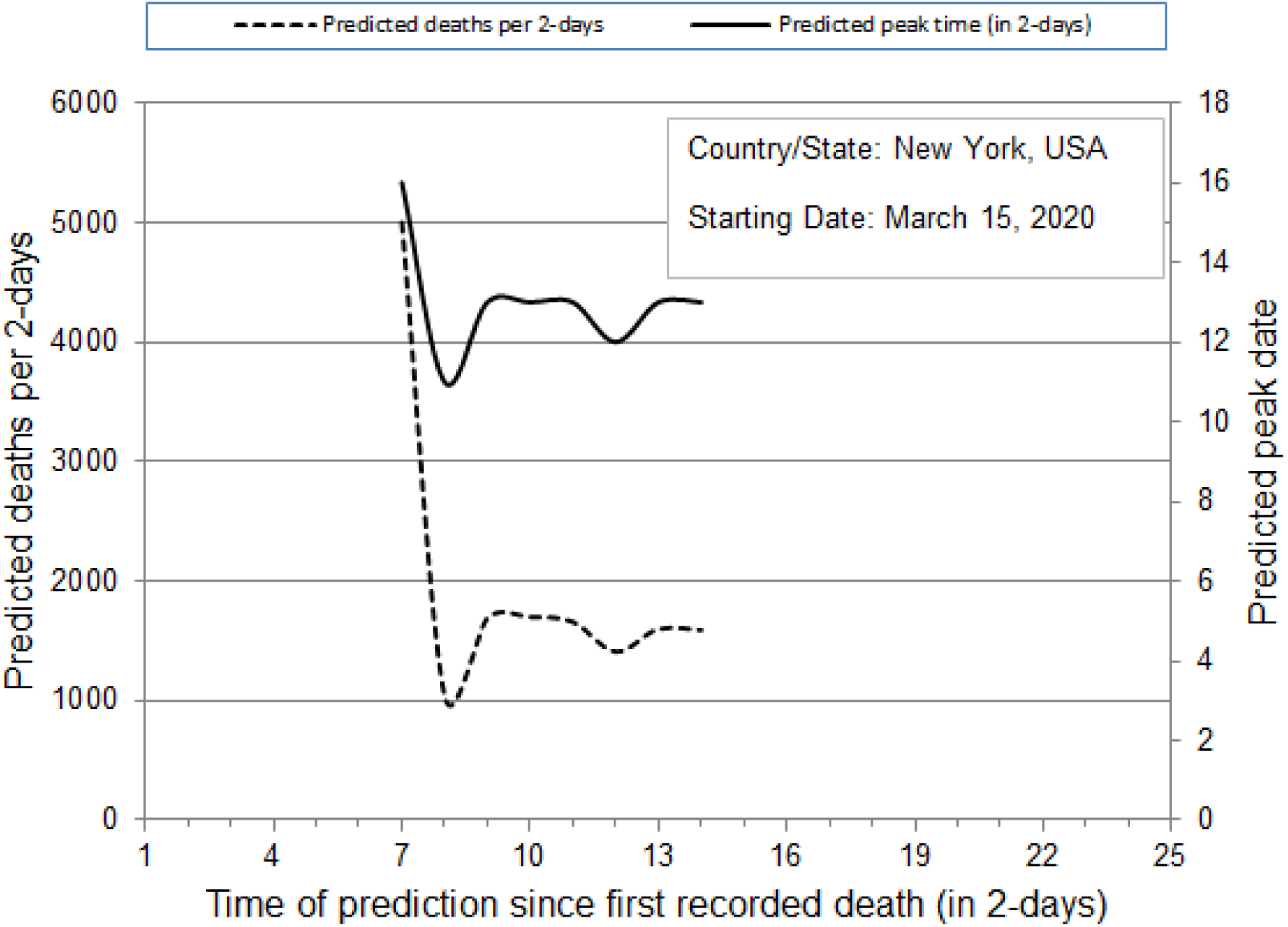
Prediction of number of deaths (dashed line), in 2-day time intervals, and peak-date of deaths (solid line) for the state of New York, USA

**Figure 6.**
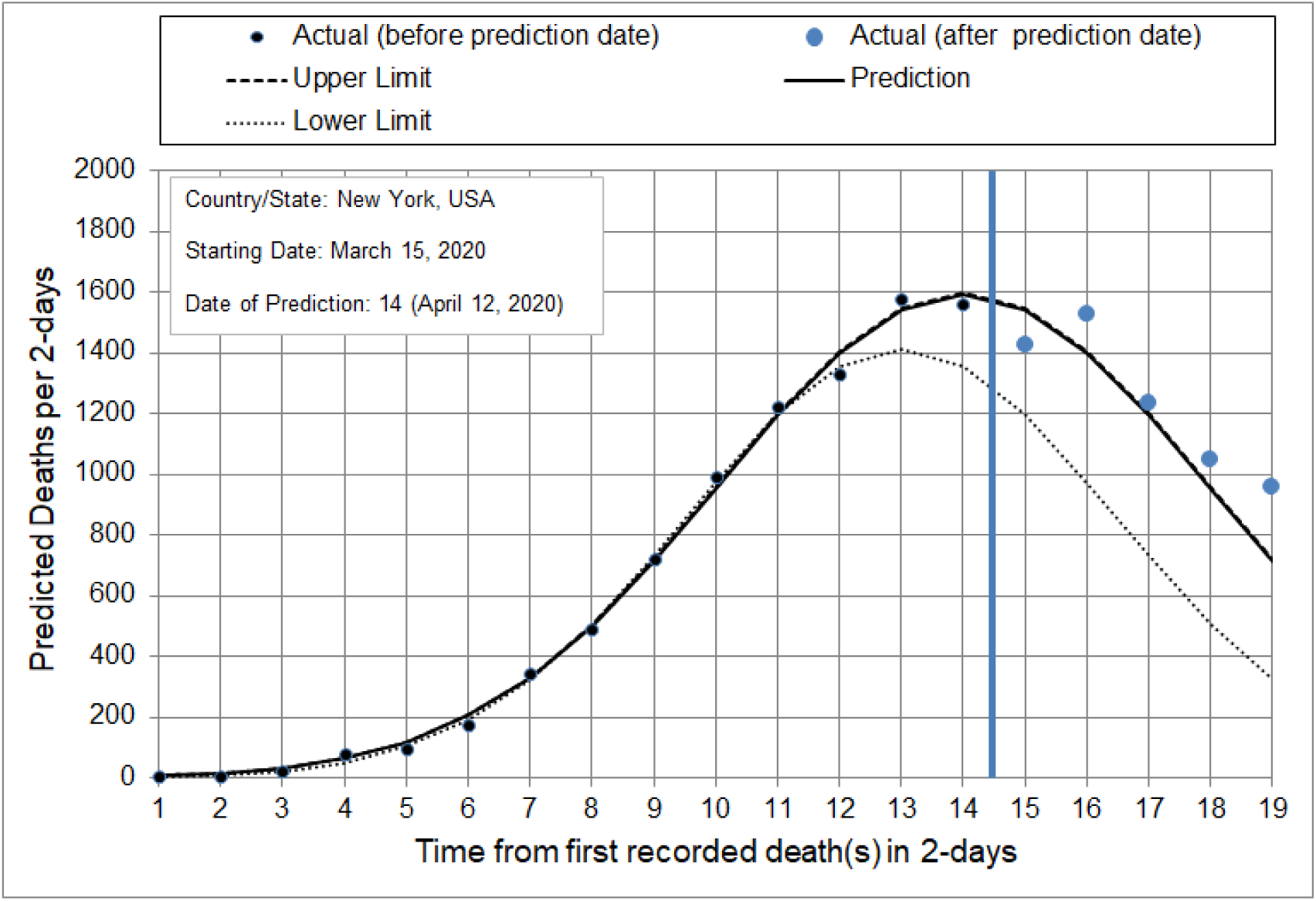
Prediction of number of deaths, in 2-day intervals, for the next ten days starting April 13, 2020, for the state of New York, USA. Black dots represent actual data until the day in which the algorithm made the prediction. Blue dots represent actual data after the day in which the algorithm made the prediction.

**Figure 7.**
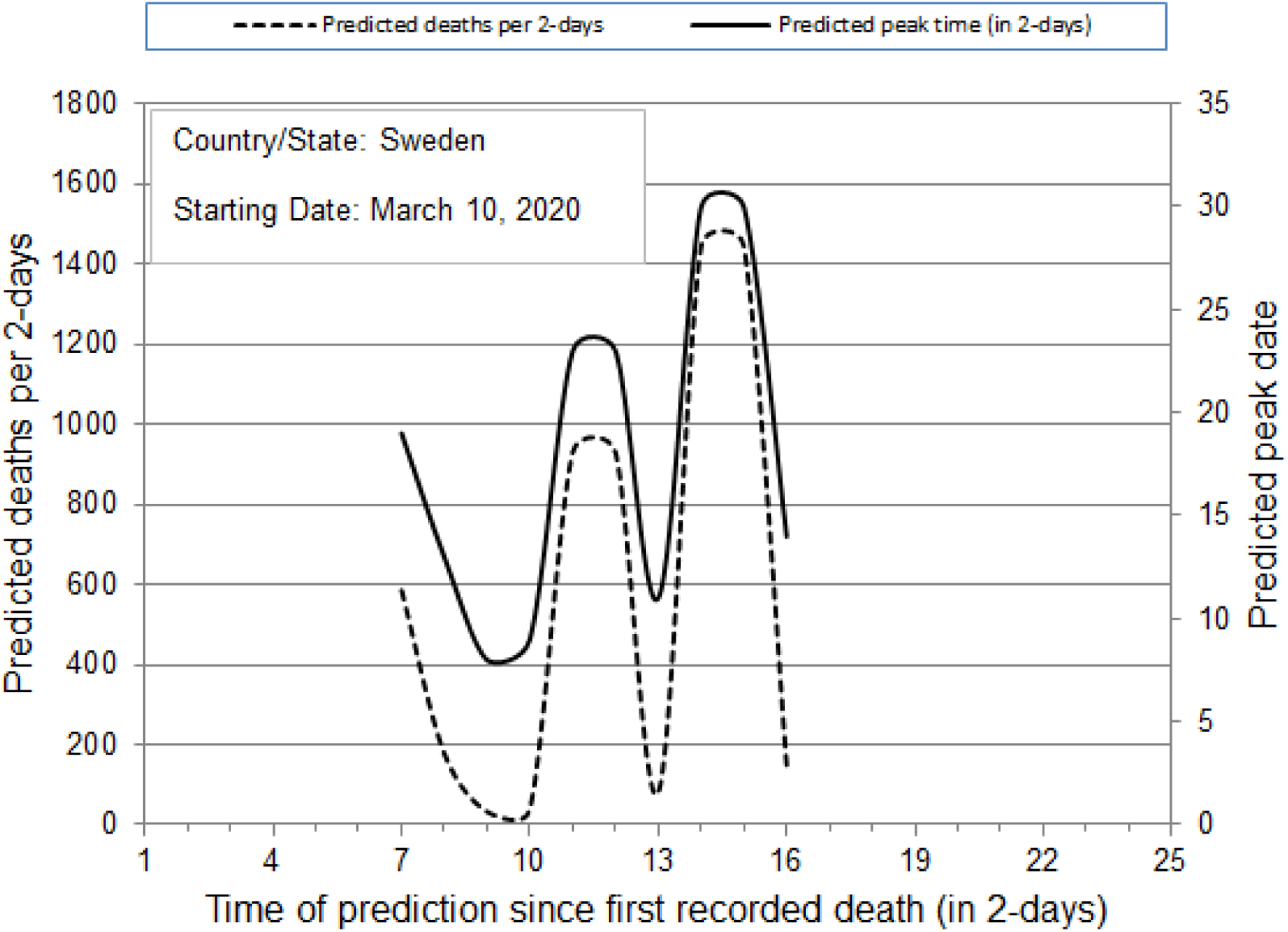
Prediction of number of deaths (dashed line), in 2-day time intervals, and peak-date of deaths (solid line) for the country of Sweden

**Figure 8.**
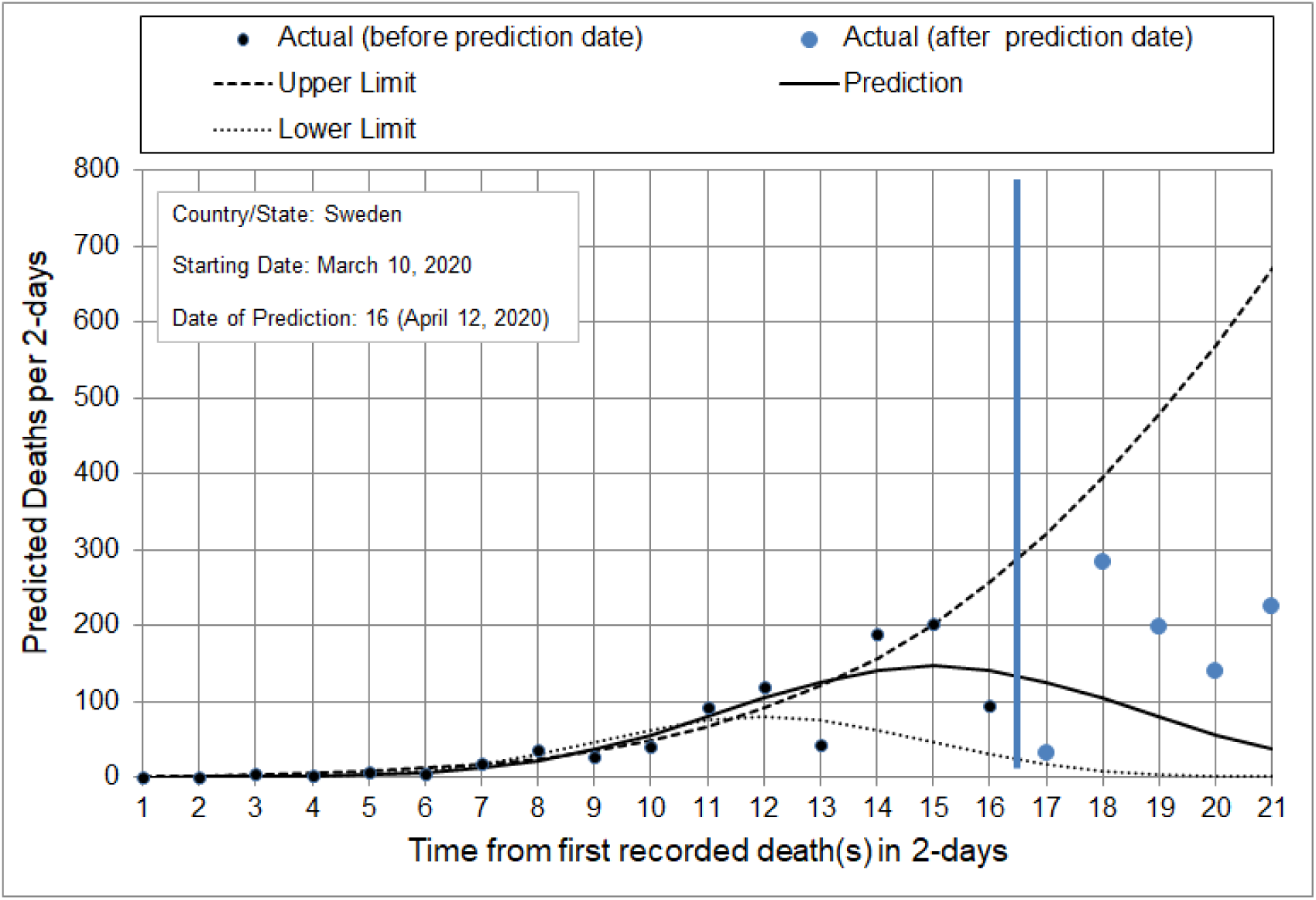
Prediction of number of deaths, in 2-day intervals, for the next ten days starting April 13, 2020, for the country of Sweden. Black dots represent actual data until the day in which the algorithm made the prediction. Blue dots represent actual data after the day in which the algorithm made the prediction.

**Figure 9.**
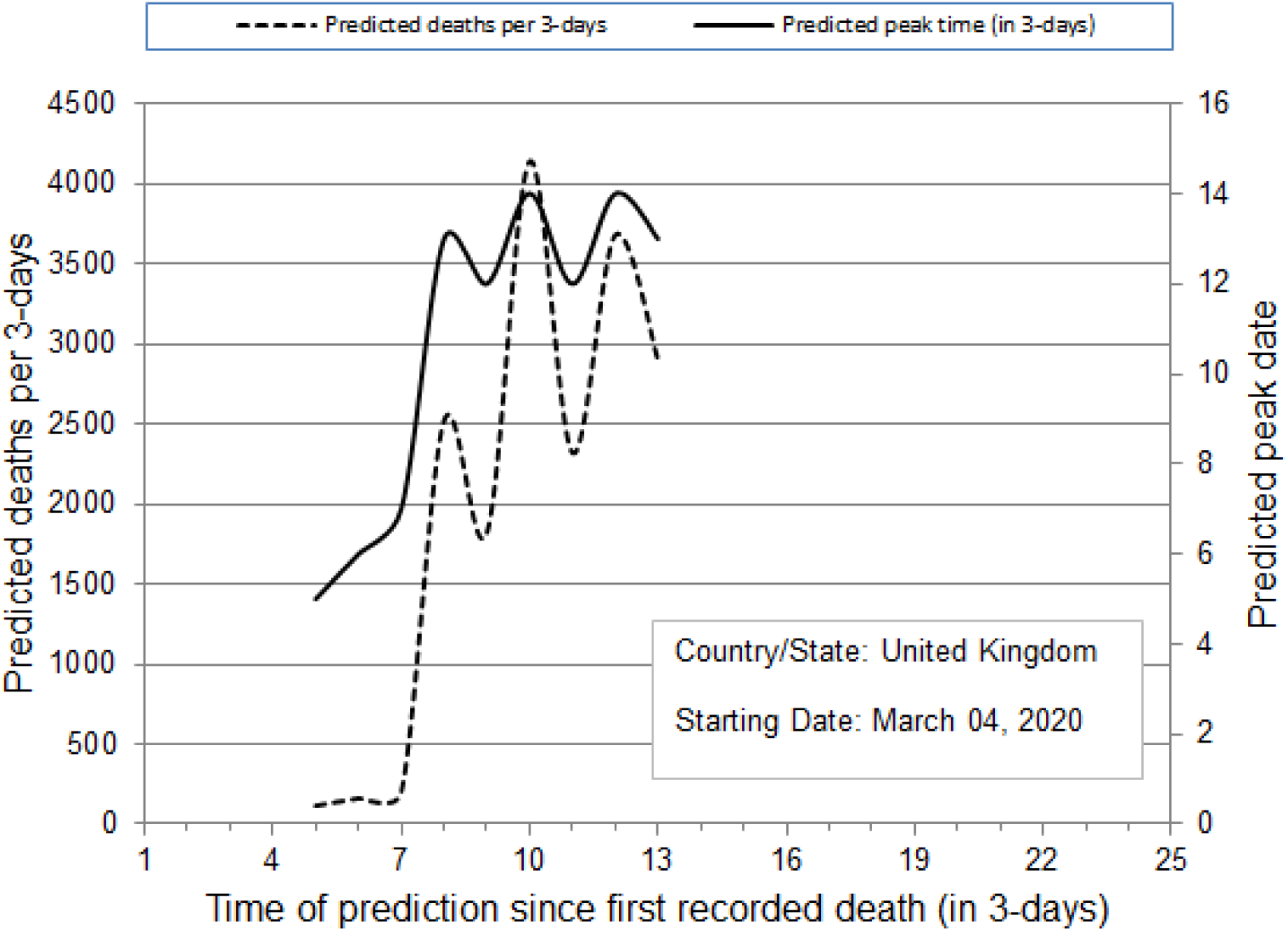
Prediction of number of deaths (dashed line), in 3-day time intervals, and peak-date of deaths (solid line) for the country of United Kingdom

**Figure 10.**
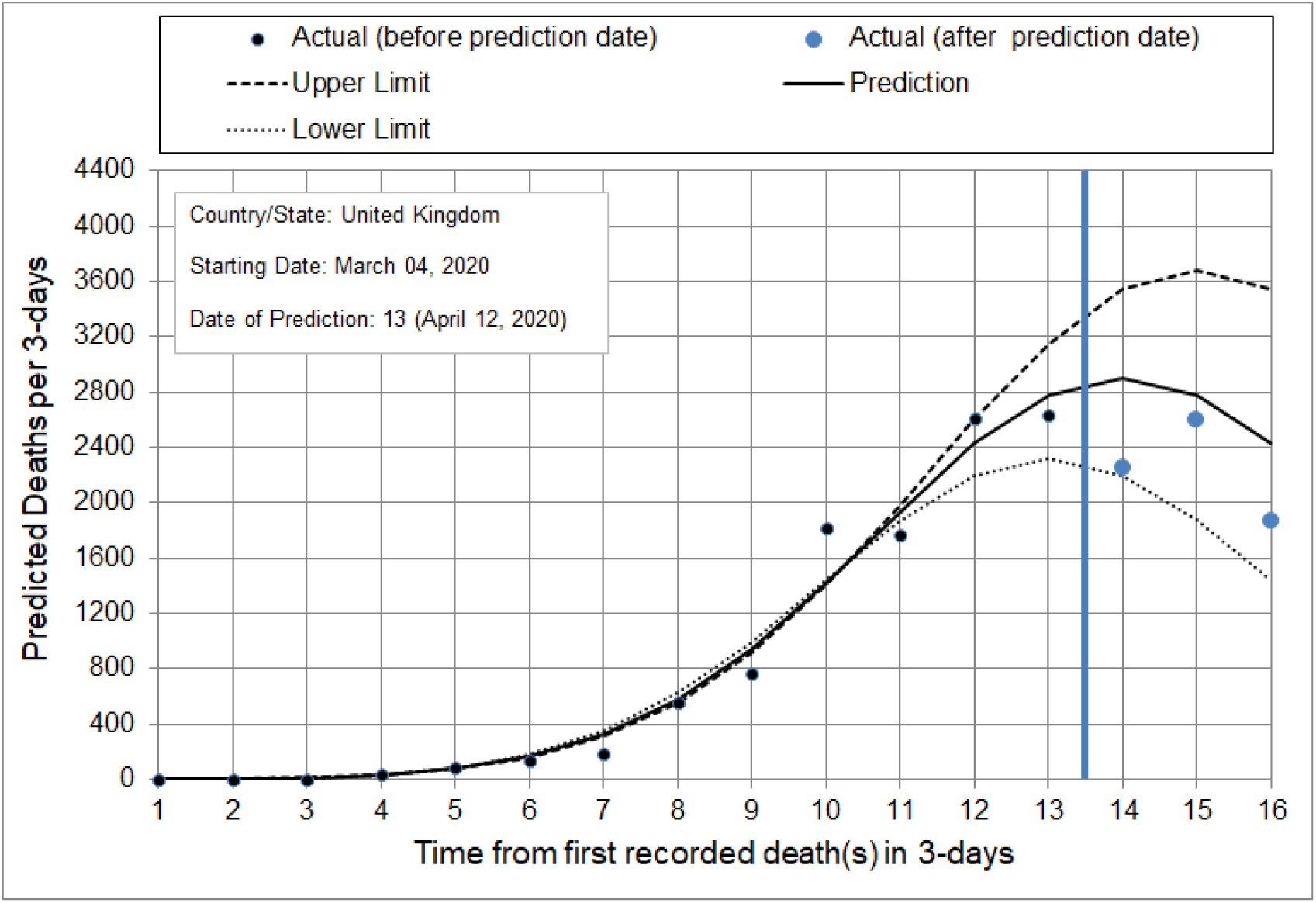
Prediction of number of deaths, in 3-day intervals, for the next ten days starting April 13, 2020, for the country of United Kingdom. Black dots represent actual data until the day in which the algorithm made the prediction. Blue dots represent actual data after the day in which the algorithm made the prediction.

**Figure 11.**
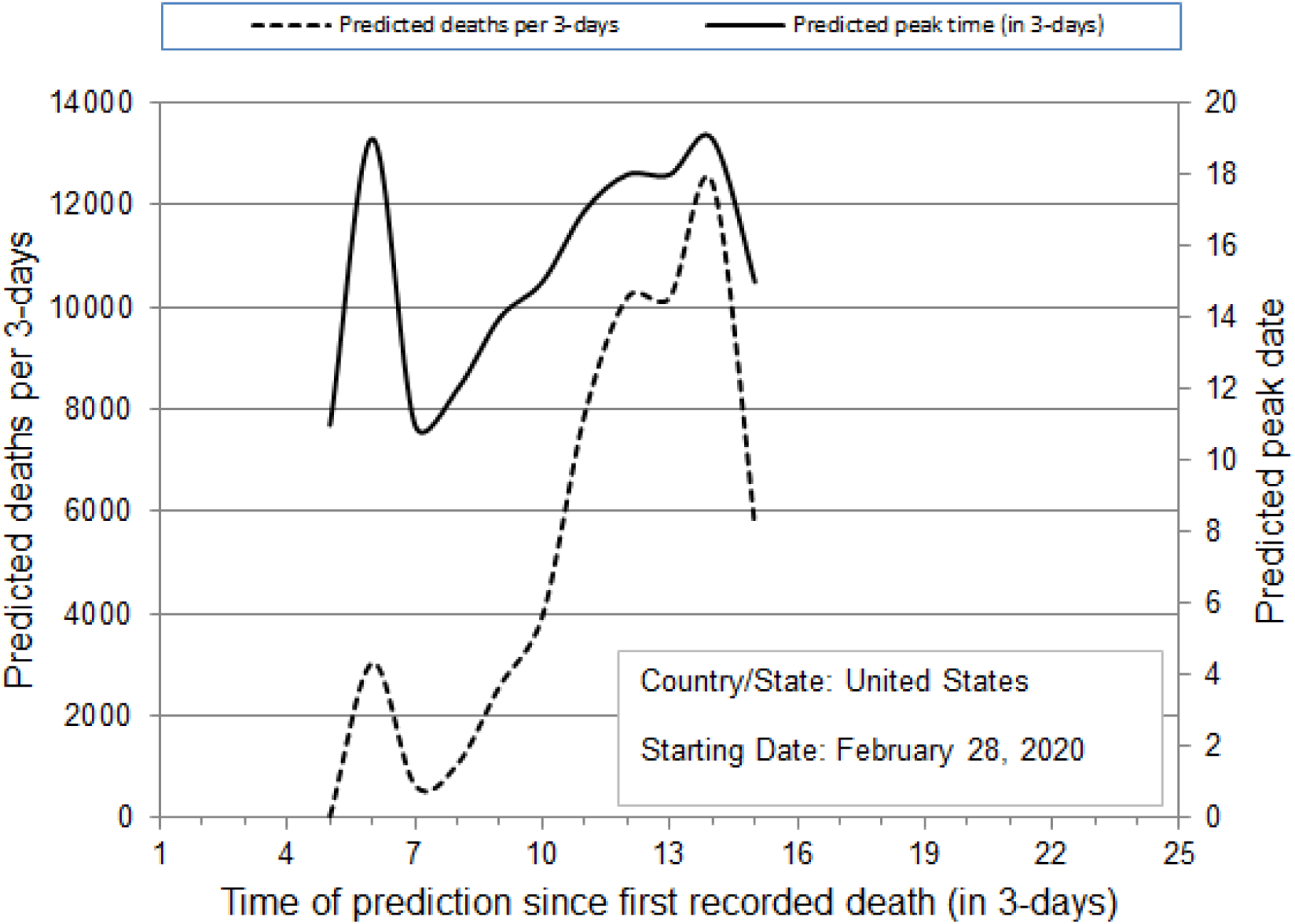
Prediction of number of deaths (dashed line), in 3-day time intervals, and peak-date of deaths (solid line) for the country of USA

**Figure 12.**
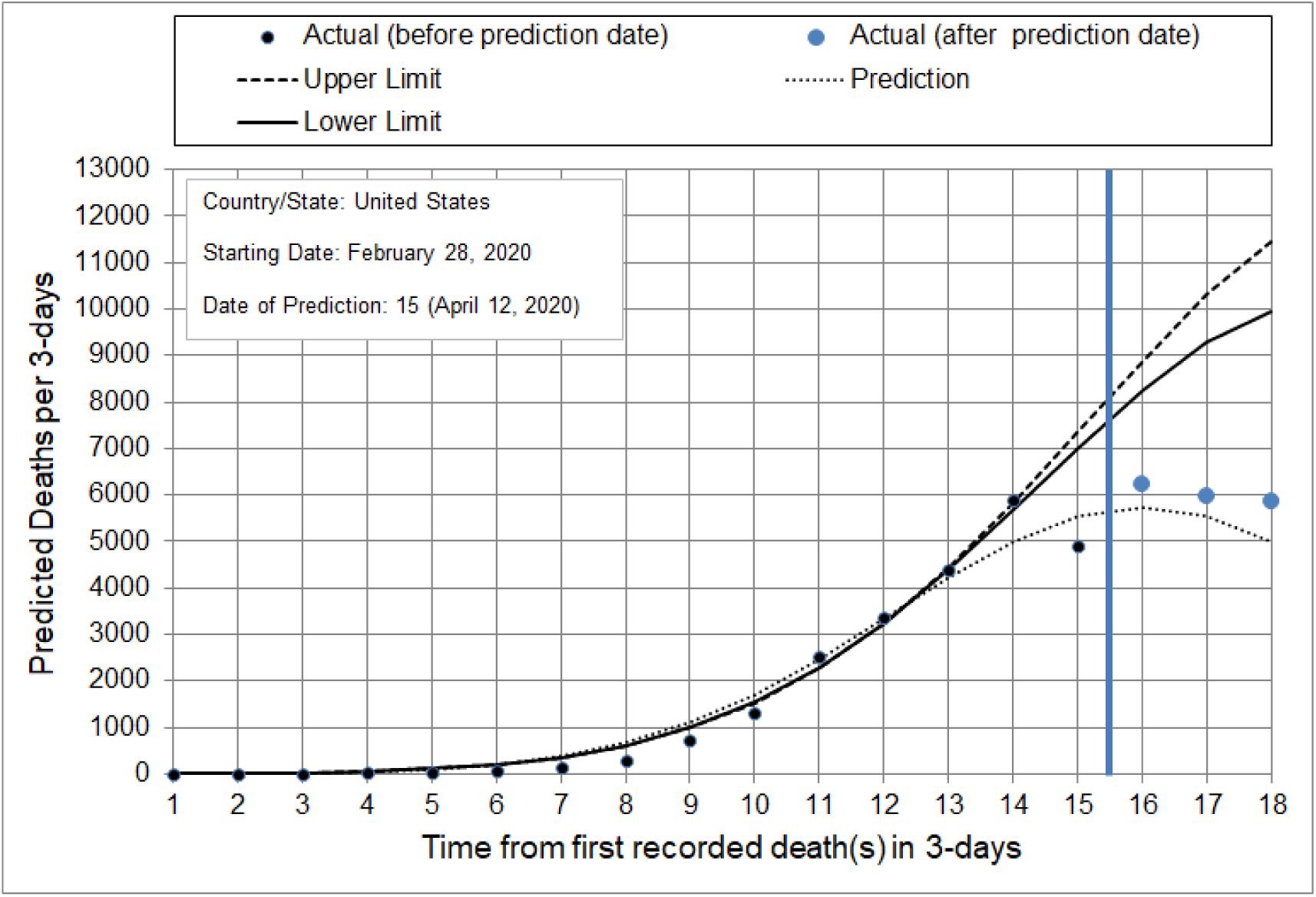
Prediction of number of deaths, in 3-day intervals, for the next ten days starting April 13, 2020, for the country of USA. Black dots represent actual data until the day in which the algorithm made the prediction. Blue dots represent actual data after the day in which the algorithm made the prediction.

## References

1. WHO Rolling updates on coronavirus disease (COVID-19): WHO characterizes COVID-19 as a pandemic. https://www.who.int/emergencies/diseases/novel-coronavirus-2019/events-as-they-happen Date accessed: March 30, 2020

2. Adam D. Modelling the Pandemic. The simulations driving the world’s response to COVID-19. Nature 580, 316–318, 2020. doi:10.1038/d41586-020-01003-6

3. Funk S, Camacho A, Kucharski AJ et al. Assessing the performance of real-time epidemic forecasts: a case-study of Ebola in the Western area region of Sierra Leone, 2014–15. PLOS Comput Biology 15(2), e1006785. doi.org/10.1371/journal.pcbi.1006785

4. Roosa K, Lee Y, Luo R, Kirpich A et al. (2020). Real-time forecasts of the COVID-19 epidemic in China from February 5th to February 24th, 2020. Infect Dis Model 5, 256–263. 10.1016/j.idm.2020.02.002

5. Shanafelt DW, Jones G, Lima M, Perrings C, Chowell G (2018). Forecasting the 2001 foot-and-mouth disease epidemic in the UK. EcoHealth. 15(2):338–347. doi:10.1007/s10393-017-1293-2

6. https://www.worldometers.info/about/

7. https://covidtracking.com/about-data

8. Burn-Murdoch J, Romei V, Giles C. Global coronavirus death toll could be 60% higher than reported. Financial Times, April 26, 2020

9. Asteris P, Douvika M, Karamani C, Skentou A, Daras T, Cavaleri L, Armaghani DJ, Chlichlia K, Zaoutis T. A novel heuristic global algorithm to predict the COVID-19 pandemic trend. MedRxiv preprint. medRxiv 2020.04.16.20068445; doi:https://doi.org/10.1101/2020.04.16.20068445

10. Ferguson NM, Laydon D, Nejati-Gilani G, Imai N et al. Report 9: impact of non-pharmaceutical interventions (NPIs) to reduce COVID-19 mortality and healthcare demand. Imperial College COVID-19 Response Team 2020

11. Yang S, Cao P, Du P, Wu Z, Zhang Z, et al. Early estimation of the case fatality rate of COVID-19 in mainland China: a data-diven analysis. Ann Transl Med 8(4), 2020.

